# A Multi-state Non-Markov Framework to Estimate Course of Chronic Disease Progression

**DOI:** 10.1101/2024.02.15.24302901

**Authors:** Ming Ding, Haiyi Chen, Feng-Chang Lin

## Abstract

In chronic disease epidemiology, the investigation of disease etiology has largely focused on one endpoint, and the progression of chronic disease as a multi-state process is understudied, representing a knowledge gap. Most existing multi-state regression models require Markov assumption and are unsuitable to describe the course of chronic disease progression that is largely non-memoryless. We propose a new non-Markov framework that allows past states to affect the transition rates of current states, and the key innovation is the conversion of a non-Markov to Markov process by conditioning on past disease history to divide disease states into substates. Specifically, we apply cause-specific Cox models, including past states as covariates, to obtain transition rates of substates, which were used to estimate transition probabilities using the discrete-time Aalen-Johansen estimator. In simulation study, the non-Markov model generated higher coverage rates of transition rates compared to Markov models, particularly for non-Markov process (By applying non-Markov and Markov models, coverage rates were 91% and 88% for Markov process with exponential distribution, 52% and 43% for Markov process with Weibull distribution, 92% and 49% for non-Markov process with exponential distribution, and 59% and 23% for non-Markov process with Weibull distribution). We applied our model to describe the course of coronary heart disease (CHD) progression, where CHD was modeled in healthy, at-risk, CHD, heart failure, and mortality states. In summary, the significance of our framework lies in the fact that transition parameters between disease sub-states may provide a more accurate description of disease course than Markov regression models and shed light on new mechanistic insight into chronic disease. Our method has the potential for wide application in chronic disease epidemiology.

## 1. INTRODUCTION

In chronic disease epidemiology, investigation of disease etiology has largely focused on one single endpoint. However, the development of chronic disease is a multi-state process, with each state playing a crucial role in affecting its progression. Understanding the course of disease progression allows us to gain new mechanistic insight into the disease at the population level. However, this field is understudied and represents a knowledge gap in epidemiology.

Most existing methods describing the progression of chronic disease are Markov models, which assume that the future state depends only on the present state and not on past history. For the Markov process, transition probabilities can be estimated parametrically from transition rates with constant or Weibull distributions using Kolmogorov differential equations,^1–4^ and nonparametrically estimated for other types of distributions using Aalen-Johansen (A-J) estimators.^5–7^ To identify factors related to the transition rate, Markov regression models have been proposed that allow the inclusion of covariates.^1, 8^ However, they are unsuitable for modeling the progression of chronic diseases that are largely non-memoryless.^9–11^ This signifies the urgent need for non-Markov methods to advance the research in estimating the course of disease progression.

Non-Markov models are a largely unexplored field, with only several nonparametric methods proposed.^12–18^ Although the A-J estimator can consistently estimate transition probabilities from time 0 to t regardless of Markov assumption,^14, 19, 20^ it is systematically biased for any two random time points in the non-Markov process.^16^ The landmark Aalen-Johansen (LMAJ) estimator accounts for non-Markov assumption and estimates transition probabilities using landmarking by dividing participants into subgroups according to the state occupation probability at a certain time point.^12^ However, the subsampling of participants lowers the power of estimation, and it is a non-parametrical method which may be difficult to incorporate covariates.^21, 22^

In this paper, we propose a new non-Markov regression framework that allows past states to affect the transition rates of current states, and the key innovation is that we divide disease states into substates to convert non-Markov to Markov process by conditioning on past disease history. Specifically, we apply cause-specific Cox models, including past states as covariates, to obtain transition rates between sub-states, which are used to obtain transition probabilities using a discrete-time A-J estimator.^8^

## 2. METHODS

### A multi-state non-Markov framework

While our method can be used for chronic disease in any number of states, we illustrate our method by assuming five states S_0_-S_4_ (**Fig 1a**). By conditioning on past states, a disease state can be divided into substates, and a non-Markov process can be converted to a Markov process (**Fig 1b**). Suppose we have survival data with disease states ascertained (**Fig 2a**), which are divided into subsets 1-4 by disease state (**Fig 2b**). Within each subset, we change the data structure from a wide form to a long-form to model transition rates with high flexibility and incorporate time-varying covariates into the framework (**Fig 2c**).

**Figure 1.**
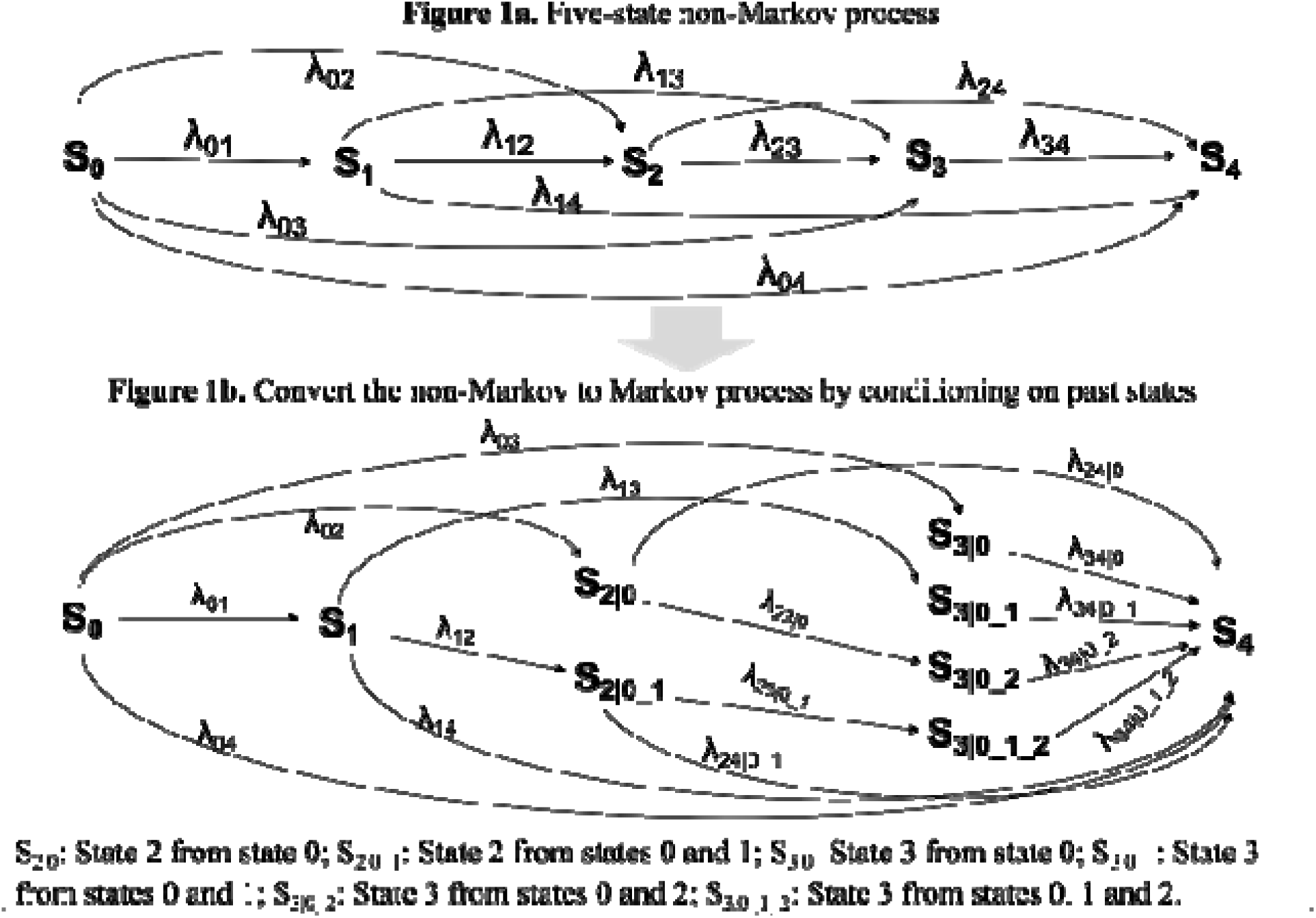
Convert non-Markov to Markov process by splitting disease states into substates.

**Figure 2.**
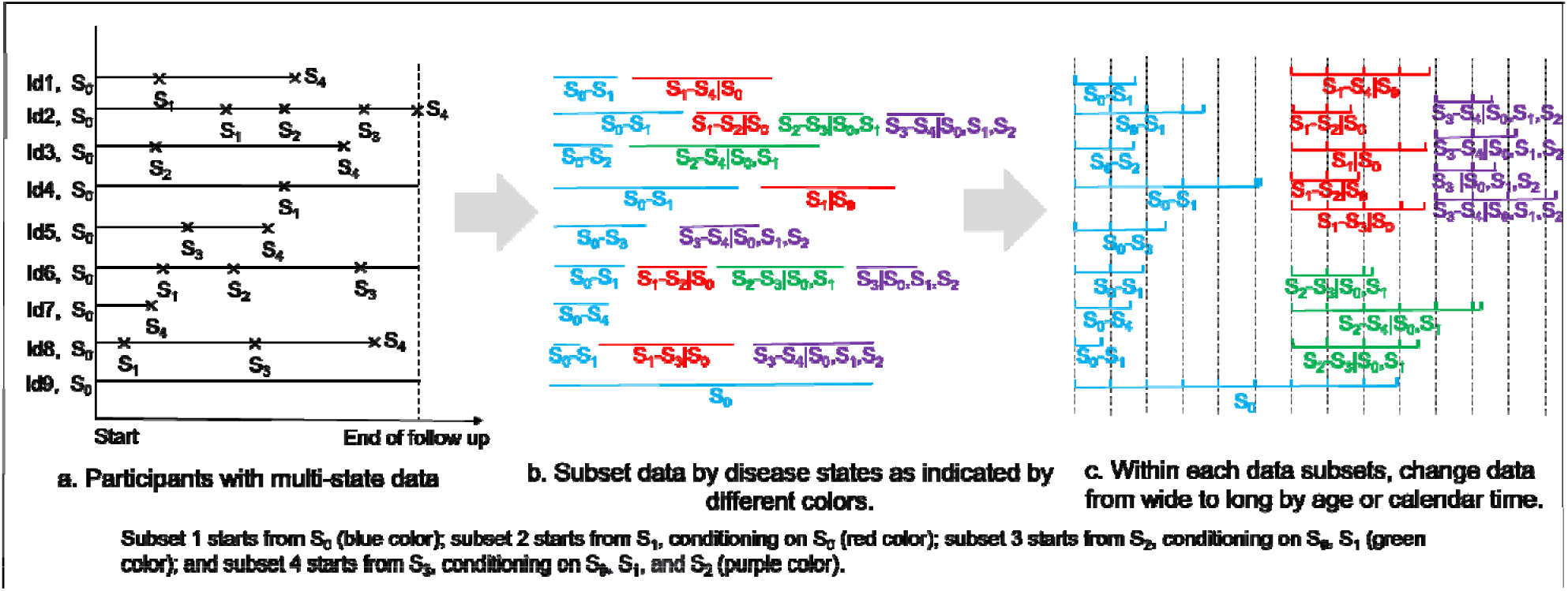
Data preparation for applying the non-Markov framework.

### 2.1. Estimate transition rate, Q(t)

Cause-specific Cox (CSC) models are used to estimate transition rates from one to multiple states. As an extension of the Cox model, CSC assumes different associations of each exposure with each specific event type.^23^ To estimate transition rates, we cut time into small intervals in changing data from wide to long-form and model time flexibly, such as using restricted cubic splines. Survival probability by the end of each interval can be estimated using the ‘predict’ command of the ‘riskRegression’ package,^24^ and hazard can be estimated as (1-survival probability)/length of the time interval. In particular, for age used as a time scale, the predicted risk at the end of each time interval is approximate to the hazard in discrete time.^24^ We fit four CSC models (**Models 1-4**) to data subsets 1-4. The influence of past history of disease on transition rates can be accounted for in models 3 and 4 by including past disease history (as well as their interaction terms with time) as covariates in the model.

**Model 1** in subset 1 starting from S_0_ state: λ_c_(t,X)=λ_0c_(t)exp(p_c_ + ∑_j_ p_jc_x_j_), where c=rate of developing S_1_, S_2_, S_3_, or S_4_ states, and x_j_ are the covariates.

**Model 2** in subset 2 starting from S_1_ state: λ_c_(t,X)=λ_0c_(t)exp(p_c_ + ∑_j_ p_jc_x_j_), where c= rate of developing S_2_, S_3_, or S_4_ states.

**Model 3** in subset 3 starting from S_2_ state: λ_c_(t,X)=λ_0c_(t)exp(p_c_ + ∑_j_ p_jc_x_j_ + p_plc_ x_pl_ + p_plc:c_x_pl_), where c=rate of developing S_3_ or S_4_ states, and x_pl_ is the past S_1_ state.

**Model 4** in subset 4 starting from S_3_ state: λ_c_(t,X)=λ_0c_(t)exp(p_c_ + ∑_j_ p_jc_x_j_ + p_plc_ x_pl_ + p_p2c_x_p2_ + p_plc:c_ x_pl_ + p_p2c:c_ x_p2_), where c=rate of developing S_4_ state, and x_pl_ and x_p2_ are past states S_1_ and S_2_, respectively.

The estimated hazards are used to construct transition rate matrix **Q(t)** (**Figure 3**), where λ_01_(t), λ_02_(t), λ_03_(t), and λ_04_(t) are the predicted risks at time t from model 1, λ_12_(t), λ_13_(t), and λ_14_(t) are predicted from model 2, λ_23|0_(t), λ_24|0_(t), λ_23|0_1_(t), and λ_24|0_1_(t) are predicted from model 3 with or without history of S_1_, λ_34|0_(t), λ_34|0_1_(t), λ_34|0_2_(t), and λ_34|0_1_2_(t) are predicted from model 4 with or without history of S_1_ and S_2_.

**Figure 3.**
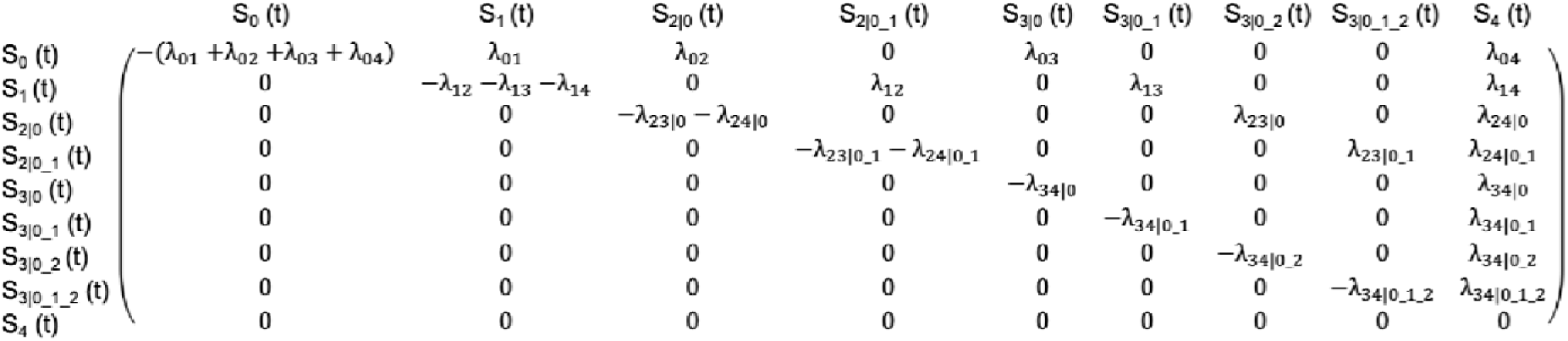
Transition rate matrix Q(t).

### 2.2. State occupational probabilities

**P(t)**, are defined as the proportion of participants occupied in each state at t and can be expressed as functions of transition rates. As P’(t) = **P**(t)* **Q**(t), **P**(t+ 1) = **P**(t) + **P**’(t) = **P**(t) + **P**(t) * **Q**(t) = **P**(t) * (19 + **Q**(t)) = (**P**(t-1) + **P**’(t-1))*(19 + **Q**(t)) = **P**(t - 1) * (*I*_9_ + **Q**(t - 1)) * (19 + **Q**(t)) = **P**(t= 0) *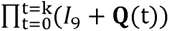, where **P**(t) = p_0_(t), p_1_ (t), p_2_|_0_(t), p_2|0_1_(t), p_3|0_(t), p_3|0_1_(t), p_3|0_2_ (t), p_3|0_1_2_(t), p_4_(t), **Q**(t) is the transition rate matrix, and *I*_9_ is an identity matrix.

### 2.3. Transition probabilities

**TP (k_1_, k_2_),** is defined as the probability of state transition over a period and can be estimated using a discrete-time A-J estimator, where TP(k_l_, k_2_)= ∏^t=k2^(I +Q(t)).

### 2.4. Use Bootstrap to obtain 95% CIs

We adopted a parametric bootstrapping approach to repeatedly draw transition rates from their corresponding distributions to estimate the variability of the parameter estimate. We repeat the resampling process 1000 times in each estimation by the CSC model. Statistically, the resulting 1000 parameter estimates represent the empirical distribution. They can be summarized with median values and 95% CI defined by 2.5th to 97.5th percentiles of the empirical distribution.

## 3. SIMULATION STUDY

### 3.1. Simulation methods

We performed a simulation study to validate the utility of the non-Markov model and compare it to the Markov model. Data were simulated in five states, and four scenarios were considered: Markov or non-Markov processes with exponential or Weibull distribution. Transition rates were simulated as constant for exponential distribution and increasing/decreasing with time for Weibull distribution. Parameters of transition rates differed by past states for the non-Markov process. We simulated survival data starting from each disease state using ‘crisk.sim’ package in R, and the parameters used for the simulation study are shown in **Table S1**. Processes with exponential distribution were a special case of Weibull distribution, with the ancillary parameters fixed at 1. We simulated the datasets 1000 times, with a sample size of 5000 for each simulation.

For each scenario, we compared the performances of the non-Markov and Markov models in the transition rates estimation of sub-states. In changing data from wide to long format, we cut time into intervals by 0.05. However, if the Cox model did not converge due to scarce cases within some intervals, we cut it by 0.1. Restricted cubic splines were used to model the event indicator highly flexibly at each time interval. In the non-Markov framework, we included past disease history and the interaction terms with time in models 3-4. Survival probability by the end of each interval was obtained, and the hazard was estimated as (1-survival probability)/length of time interval. We estimated transition rates at time points 0.3, 0.6, 0.9, 1.2, and 1.5 when most simulated participants developed the endpoint. The estimated transition rates were compared to theoretical rates, calculated as

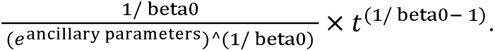

Mean squared error (MSE), coverage, and width of 95% CI were obtained. MSE is the average squared difference between the estimate and true value. Coverage is defined as the proportion of 95% CIs covering the true value over all simulated datasets. We compared the coverage to determine which method was closer to the 95% nominal coverage rate. Width of 95% CI is the difference between 95% upper and lower levels. To compare transition rates across models in a summarized manner, we calculated mean MSE, coverage, and width of 95% CI across all states and all time points.

### 3.2. Simulation results

In Markov scenarios, both Markov and non-Markov models worked well, as expected (**Table 1**). In non-Markov scenarios, the non-Markov model outperformed the Markov model by showing lower MSE and higher coverage rates. For a non-Markov process with exponential distribution, the coverage rates were 92% and 49%, respectively, using non-Markov and Markov models. For a non-Markov process with Weibull distribution, the coverage rates were 59% and 23% using the non-Markov and Markov models, respectively. The better performance of non-Markov compared to the Markov model was mainly due to the lower MSE and higher coverage rates in estimated transition rates from states 2 and 3 and transition rates from states 3 to 4, which were affected by past disease history (**Table 2**).

**Table 1.** A simulation study estimating transition rates (%) compared the non-Markov regression model to Markov model under different scenarios.

|  | Markov process with exponential distribution |  | Markov process with Weibull distribution |  |
| --- | --- | --- | --- | --- |
|  | Cox Markov model | Non-Markov model | Cox Markov model | Non-Markov model |
| MSE | 8.88 | 16.80 | 60.30 | 59.46 |
| Width of 95% CI | 8.85 | 13.12 | 8.93 | 11.88 |
| Coverage rate | 88% | 91% | 43% | 52% |
|  | Non-Markov process with exponential distribution |  | Non-Markov process with Weibull distribution |  |
|  | Cox Markov model | Non-Markov model | Cox Markov model | Non-Markov model |
| MSE | 482.94 | 17.37 | 420.89 | 47.52 |
| Width of 95% CI | 6.63 | 11.25 | 5.07 | 10.16 |
| Coverage rate | 49% | 92% | 23% | 59% |

Mean squared error (MSE) is defined as the average squared difference between estimated value and true value. Width of 95% confidence interval (CI) is defined as the difference between 95% upper and lower levels. Coverage is defined as the proportion of 95% CIs including the true value over all simulated datasets.

**Table 2.**
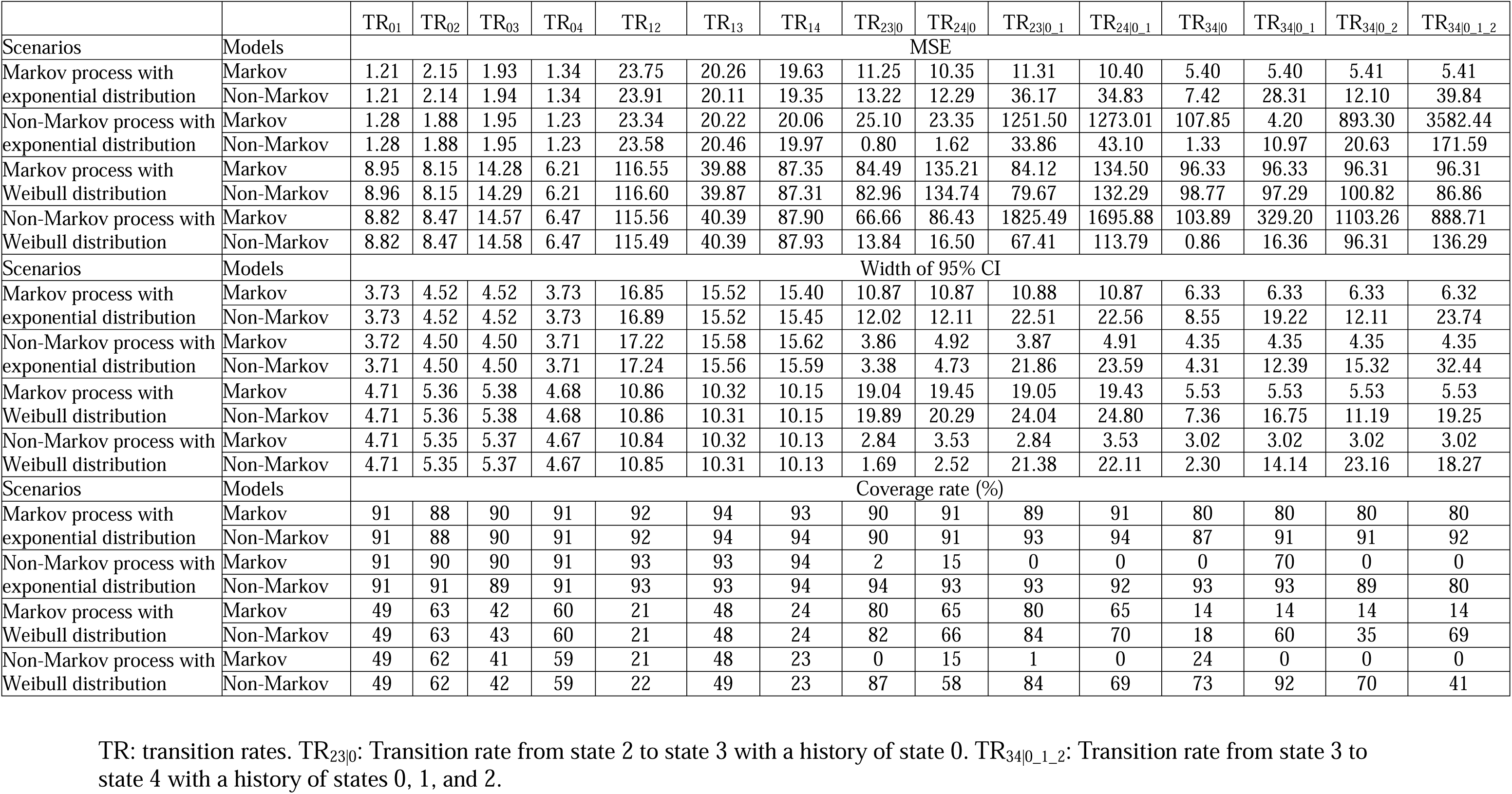
A simulation study estimating transition rates (%) by disease substates compared the non-Markov regression model to Markov model under different scenarios.

## 4. APPLICATION STUDY

### 4.1. Study population

The Atherosclerosis Risk in Communities Study (ARIC) study was designed to investigate the causes of atherosclerosis and its clinical outcomes, as well as variations in cardiovascular risk factors and disease by sex and race.^25^ The enrolled participants ARIC underwent a phone interview and clinic visit at baseline and were followed up by telephone calls and re-examinations. Participants were contacted periodically by phone and interviewed about interim hospital admissions, cardiovascular outpatient diagnoses, and deaths. Participants who reported CVD-related events were asked to provide medical records that were reviewed by physicians.^26^ For each event, the month and year of diagnosis were recorded as the diagnosis date. Heart failure was ascertained by surveillance calls or clinic visits and was verified from death certificates, medical records, and outpatient diagnoses. Deaths were identified from systematic searches of the vital records of states and of the National Death Index, supplemented by reports from family members and postal authorities.^27^ We obtained ARIC data through BioLINCC, an open repository created by NHLBI, with the IRB deemed exempt by the University of North Carolina (UNC)-Chapel Hill Review Board.

### 4.2. Methods

We modeled coronary heart disease (CHD) progression in five states: Healthy, at risk, CHD, heart failure, and mortality. At-risk state was defined as development of hypertension, hyperlipidemia, or diabetes. Hypertension was defined as blood pressure ≥140/90 mmHg or a history of hypertension or use of blood pressure medications.^28^ Hyperlipidemia included primary hypertriglyceridemia (≥175 mg/dL) or primary hypercholesterolemia (LDL-c 160–189 mg/dL, and/or non-HDL-c 190–219 mg/dL).^29^ Diabetes was defined as fasting glucose ≥7.7mmol/L.^30^ We calculated the time of incidence of at-risk state as the earliest time that any of the risk factors were documented.

We assume a forward model of CHD progression while recognizing that it can be backward (*e.g.*, some participants may develop CHD before at-risk state). Our model can deal with this reverse scenario by treating CHD to at-risk state as a new transition. However, given the low proportion of participants who develop CHD before the at-risk state, including this transition may lower study power. Instead, we classify those following reverse transition to a path of forward transition (*e.g.*, ‘healthy → CHD → at risk → mortality’ is classified to the path ‘healthy → CHD → mortality’).

We applied our non-Markov framework to examine the dynamics of CHD progression. Briefly, we divided the longitudinal data into four subsets, “individuals start from healthy state to incidence of at-risk, CHD, heart failure, or mortality, whichever outcome came first,” “individuals start from at-risk state to incidence of CHD, heart failure, or mortality,” “individuals start from CHD state to incidence of heart failure or mortality,” and “individuals start from heart failure state to incidence of mortality.” Within each subset, we changed the data into a long format by cutting them into small intervals by age. We modeled age flexibly in cubic terms and included past disease history and the interaction terms with age as covariates in models 3 and 4.

### 4.3. Results

Our study included 15,027 participants who did not develop CHD or heart failure at baseline. During a median of 27 years of follow-up, our study documented 13,043 at-risk participants, 2565 incident cases of CHD, 3283 incident cases of heart failure, and 7677 incident cases of mortality. The transition rates from healthy, at-risk, or CHD states to subsequent states were low at mid-age and gradually increased with age (**Figure 4**). There was a drastic increase in the transition rate from healthy or at-risk status to mortality after 80 years. The transition rate from heart failure to mortality was higher across all ages compared to transition rates between other states. For the transition from heart failure to mortality, participants with a disease history had a higher rate of mortality than those without, with the highest transition rate observed among participants with a history of risk factors and CHD. We provided transition rates between substates at ages 70 and 90 years in **Table 3**.

**Figure 4.**
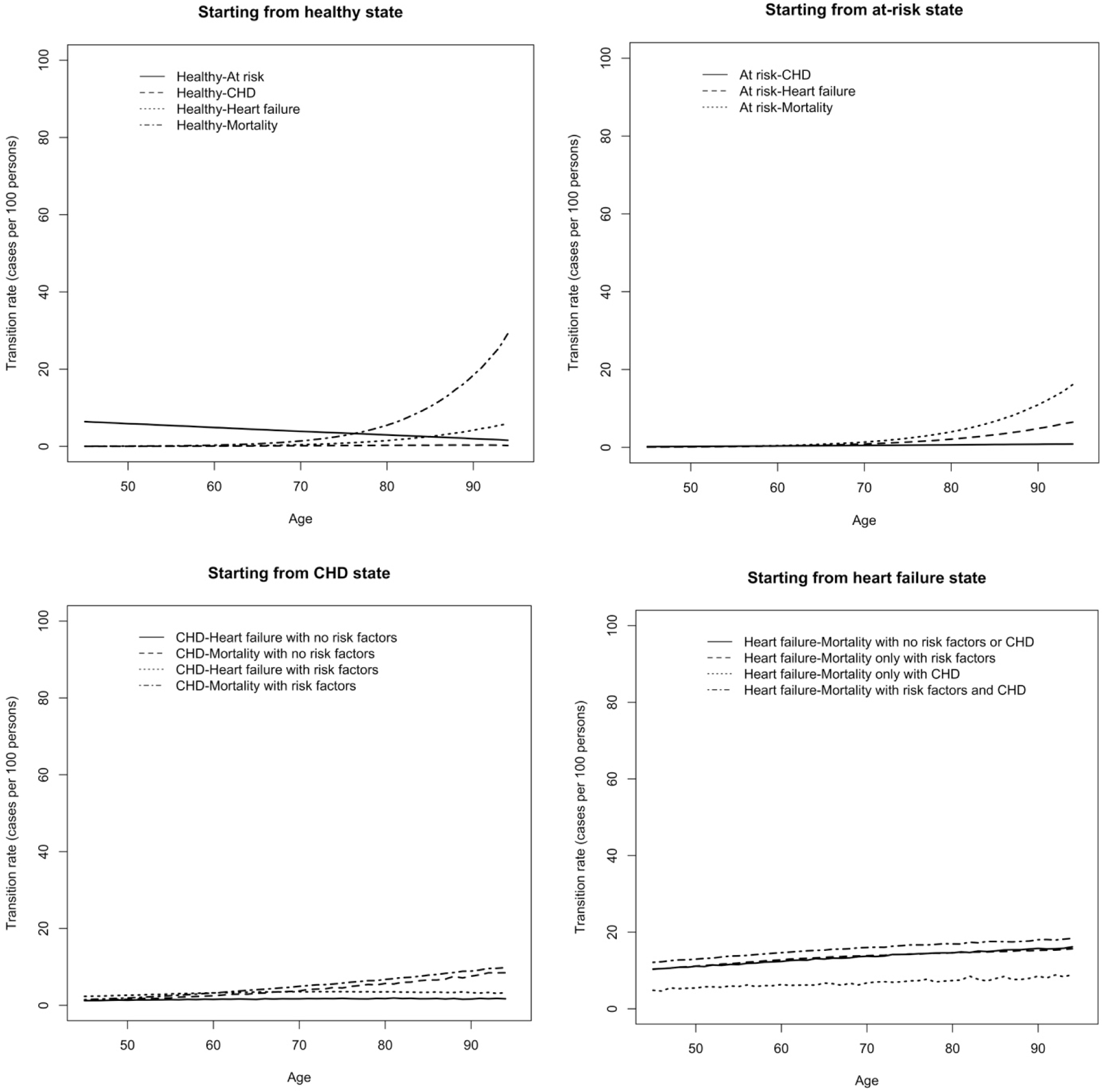
Age-specific transition rates (cases per 100 persons) from age 45 to 94 years estimated using the non-Markov framework in the Atherosclerosis Risk in Communities Study (ARIC) study.

**Table 3.** Transition rates (cases per 100 persons) at age 70 and 90 years estimated using the non-Markov framework in the Atherosclerosis Risk in Communities Study (ARIC) study.

|  | Age 70 years | Age 90 years |
| --- | --- | --- |
| **Transition from healthy state** |  |  |
| Healthy → at-risk | 4.21 (3.99, 4.41) | 2.39 (2.12, 2.66) |
| Healthy → CHD | 0.21 (0.17, 0.26) | 0.75 (0.34, 1.15) |
| Healthy → heart failure | 0.45 (0.38, 0.52) | 5.13 (3.78, 6.52) |
| Healthy → mortality | 1.34 (1.22, 1.47) | 19.12 (16.39, 21.63) |
| **Transition from at-risk state** |  |  |
| At-risk → CHD | 0.51 (0.48, 0.54) | 0.91 (0.78, 1.03) |
| At-risk → heart failure | 0.83 (0.80, 0.88) | 4.85 (4.46, 5.27) |
| At-risk → mortality | 1.28 (1.23, 1.33) | 10.58 (10.00, 11.25) |
| **Transition from CHD state with no risk factors** |  |  |
| CHD → heart failure | 2.81 (1.69, 3.94) | 3.42 (1.58, 5.24) |
| CHD → mortality | 5.25 (3.60, 6.84) | 12.02 (7.14, 16.51) |
| **Transition from CHD state with risk factors** |  |  |
| CHD → heart failure | 3.99 (3.51, 4.43) | 4.87 (3.49, 6.41) |
| CHD → mortality | 5.27 (4.72, 5.77) | 11.96 (8.87, 15.26) |
| **Transition from heart failure state with no risk factors or CHD** |  |  |
| Heart failure → mortality | 15.80 (13.59, 17.91) | 19.33 (15.61, 23.06) |
| **Transition from heart failure state only with risk factors** |  |  |
| Heart failure → mortality | 14.60 (13.78, 15.45) | 17.82 (15.12, 20.32) |
| **Transition from heart failure state only with CHD** |  |  |
| Heart failure → mortality | 13.43 (6.21, 20.91) | 16.96 (7.96, 25.24) |
| **Transition from heart failure state with risk factors and CHD** |  |  |
| Heart failure → mortality | 17.65 (15.91, 19.61) | 21.54 (17.67, 25.02) |

We estimated the transition probability from each state to the following states starting from 45 years old (**Figure 5**). Participants who started from a healthy state were very likely to develop at-risk state over follow-up. The transition probability from healthy to at-risk state first increased and then decreased due to participants’ transition to next states. The risk of mortality was higher for participants who had CHD at 45 years old with a history of risk factors than those without. For participants who had heart failure at 45 years old, the risk of mortality was greater than 90% at age 70 years without disease history and even higher for those with a history of risk factors and CHD. We provided transition probabilities between substates at ages 70 and 90 years in **Table 4**.

**Figure 5.**
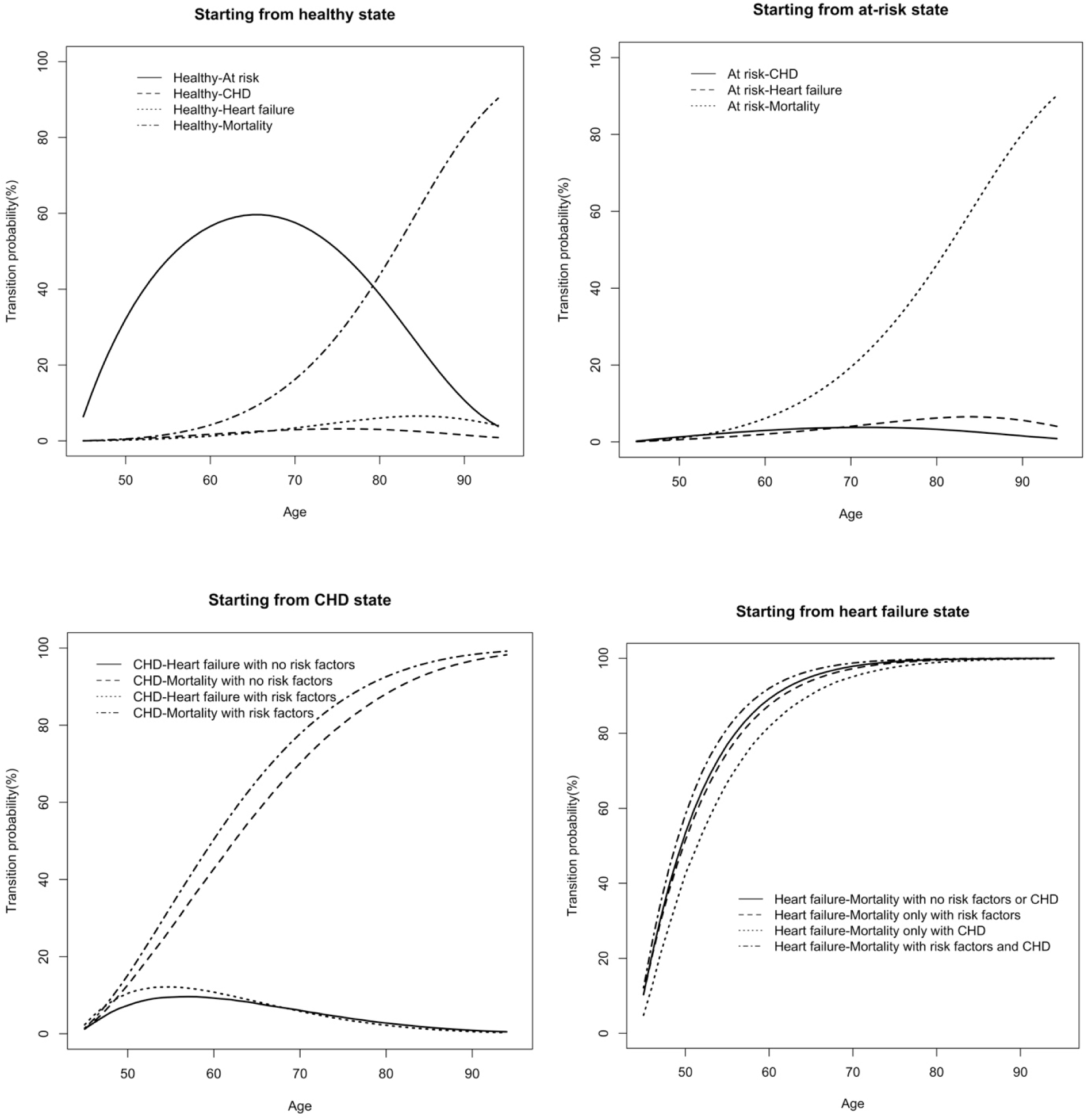
Transition probabilities (%) starting from age 45 years estimated using the non-Markov framework in the Atherosclerosis Risk in Communities Study (ARIC) study.

**Table 4.** Transition probabilities (%) from age 45 years to age 70 and 90 years estimated using the non-Markov framework in the Atherosclerosis Risk in Communities Study (ARIC) study.

|  | Age 70 years | Age 90 years |
| --- | --- | --- |
| **Transition from healthy state** |  |  |
| Healthy → Healthy | 20.68 (20.35, 21.00) | 1.17 (1.09, 1.25) |
| Healthy → at risk | 58.67 (58.37, 58.96) | 13.28 (13.05, 13.52) |
| Healthy → CHD | 2.91 (2.85, 2.98) | 1.85 (1.72, 1.97) |
| Healthy → heart failure | 3.20 (3.13, 3.26) | 6.22 (5.97, 6.50) |
| Healthy → mortality | 14.54 (14.40, 14.69) | 77.48 (77.11, 77.82) |
| **Transition from at-risk state** |  |  |
| At risk → At risk | 74.50 (74.32, 74.71) | 14.34 (14.08, 14.60) |
| At risk → CHD | 3.84 (3.75, 3.92) | 1.89 (1.74, 2.04) |
| At risk → heart failure | 3.91 (3.83, 3.98) | 6.17 (5.88, 6.48) |
| At risk → mortality | 17.76 (17.59, 17.90) | 77.61 (77.21, 77.97) |
| **Transition from CHD state with no risk factors** |  |  |
| Remain in CHD state | 22.39 (20.75, 24.06) | 1.90 (1.58, 2.28) |
| CHD → heart failure | 7.69 (6.42, 9.21) | 1.31 (1.02, 1.71) |
| CHD → mortality | 69.90 (67.81, 72.06) | 96.78 (96.20, 97.27) |
| **Transition from CHD state with risk factors** |  |  |
| Remain in CHD state | 17.10 (16.52, 17.71) | 1.10 (0.97, 1.23) |
| CHD → heart failure | 6.59 (6.30, 6.93) | 0.73 (0.66, 0.81) |
| CHD → mortality | 76.30 (75.62, 76.99) | 98.17 (98.01, 98.32) |
| **Transition from heart failure state with no risk factors** |  |  |
| Remain in heart failure state | 2.21 (1.96, 2.52) | 0.05 (0.04, 0.06) |
| Heart failure → mortality | 97.79 (97.48, 98.04) | 99.95 (99.94, 99.96) |
| **Transition from heart failure state only with risk factors** |  |  |
| Remain in heart failure state | 3.08 (2.95, 3.23) | 0.09 (0.08, 0.10) |
| Heart failure → mortality | 96.92 (96.77, 97.05) | 99.91 (99.90, 99.92) |
| **Transition from heart failure state only with CHD** |  |  |
| Remain in heart failure state | 3.96 (2.79, 5.59) | 0.14 (0.09, 0.25) |
| Heart failure → mortality | 96.04 (94.41, 97.21) | 99.86 (99.75, 99.91) |
| **Transition from heart failure state with risk factors and CHD** |  |  |
| Remain in heart failure state | 1.40 (1.27, 1.53) | 0.02 (0.02, 0.02) |
| Heart failure → mortality | 98.60 (98.47, 98.73) | 99.98 (99.98, 99.98) |

We estimated state occupational probability for participants starting from healthy states at age 45. The proportion of participants in the healthy state decreased over time, and the proportion in the mortality state increased over time (**Figure 6**). The proportion of at-risk participants first increased and then decreased because these participants moved on to the next states, such as CHD, heart failure, or mortality states. At each age, the proportions of participants in CHD or heart failure states were low, either due to their low incidences from previous states or their high risks of moving to the next states. For example, the proportion of participants who were in healthy, at-risk, CHD, heart failure, and mortality states at age 90 years were 1.17, 13.28, 1.85, 6.12, and 77.47%, respectively (**Table 5**). Most of the participants in CHD or heart failure states had a history of risk factors: 87% of CHD participants had risk factors, and 82% of heart failure participants had risk factors or CHD history.

**Figure 6.**
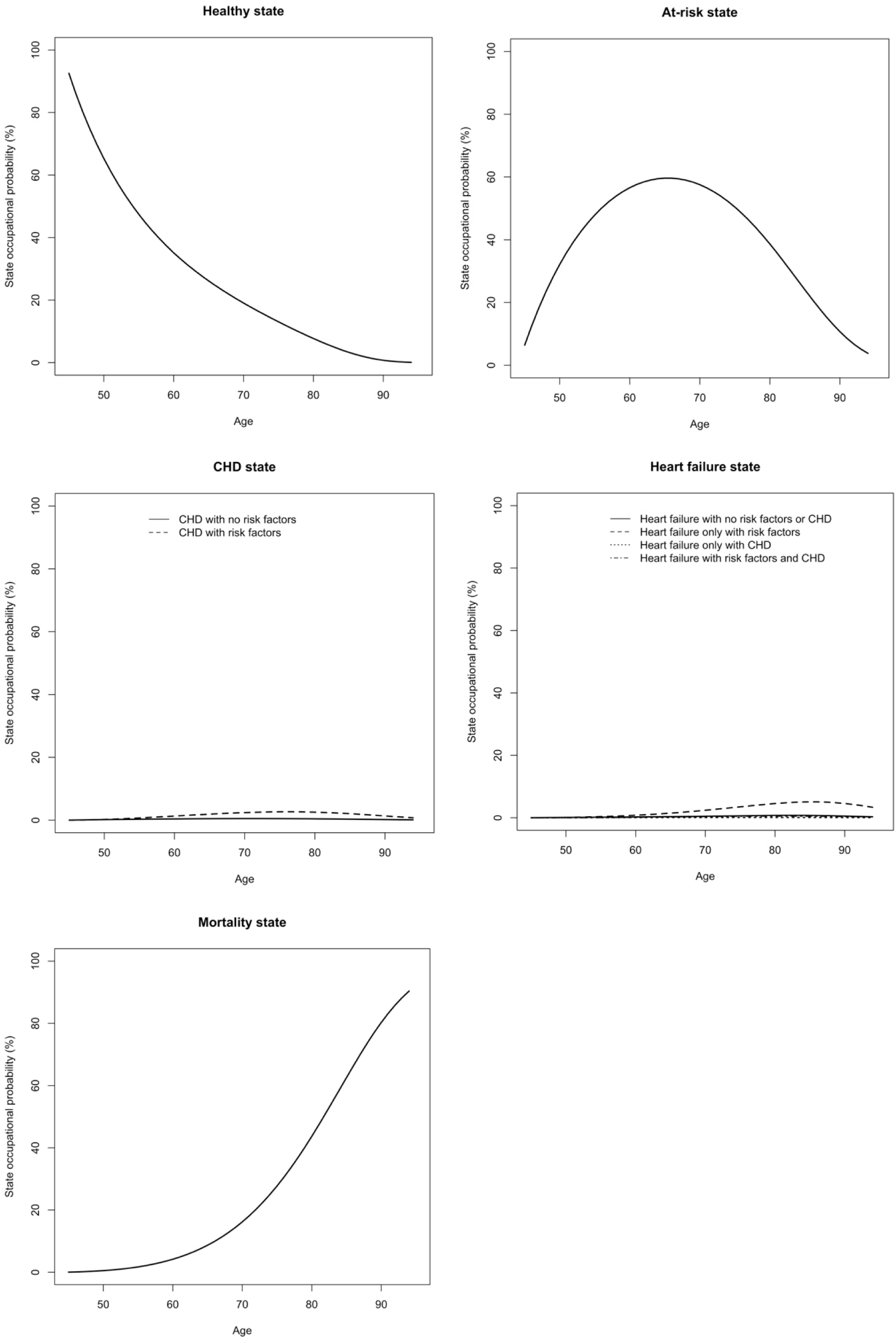
State occupational probabilities (%) starting from age 45 years estimated using the non-Markov framework in the Atherosclerosis Risk in Communities Study (ARIC) study.

**Table 5.**
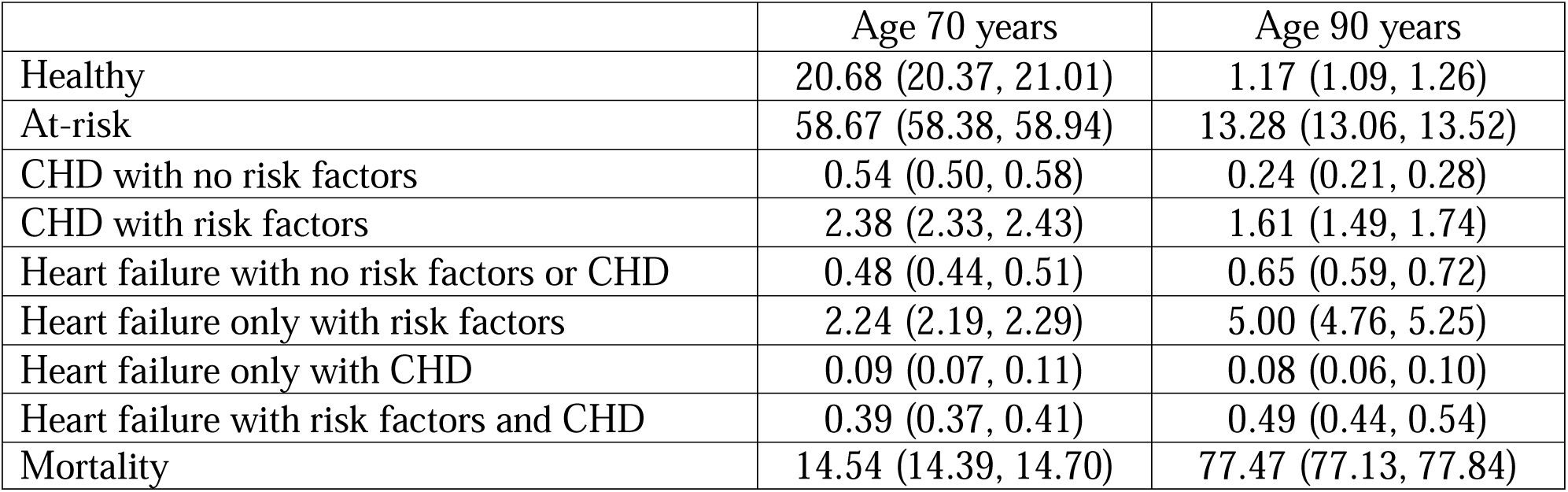
State occupational probabilities (%) from age 45 years to age 70 and 90 years estimated using the non-Markov framework in the Atherosclerosis Risk in Communities Study (ARIC) study.

|  | Age 70 years | Age 90 years |
| --- | --- | --- |
| Healthy | 20.68 (20.37, 21.01) | 1.17 (1.09, 1.26) |
| At-risk | 58.67 (58.38, 58.94) | 13.28 (13.06, 13.52) |
| CHD with no risk factors | 0.54 (0.50, 0.58) | 0.24 (0.21, 0.28) |
| CHD with risk factors | 2.38 (2.33, 2.43) | 1.61 (1.49, 1.74) |
| Heart failure with no risk factors or CHD | 0.48 (0.44, 0.51) | 0.65 (0.59, 0.72) |
| Heart failure only with risk factors | 2.24 (2.19, 2.29) | 5.00 (4.76, 5.25) |
| Heart failure only with CHD | 0.09 (0.07, 0.11) | 0.08 (0.06, 0.10) |
| Heart failure with risk factors and CHD | 0.39 (0.37, 0.41) | 0.49 (0.44, 0.54) |
| Mortality | 14.54 (14.39, 14.70) | 77.47 (77.13, 77.84) |

## 5. DISCUSSION

We proposed a new non-Markov regression model that allows past states to affect current states, and the innovation lies in that a non-Markov process is converted to a Markov process by conditioning on past states and dividing disease states into substates. The simulation study showed that our method generated less biased transition rates for the non-Markov process compared to Markov regression models, which was observed for both exponential and Weibull distributions. In the application study, we applied our model to describe the course of CHD progression.

As the leading cause of mortality in the U.S.,^31^ the etiology and prevention of CHD have been investigated for decades and studies have demonstrated that CHD is largely preventable, with modifiable risk factors accounting for 82% of CHD events,^32^ including smoking,^33^ body mass index (BMI),^34–36^ diet quality,^37–39^ and physical activity.^40, 41^ However, these studies largely focused on one single endpoint, such as incidences of hypertension, CHD, or heart failure.^42–47^ A paucity of studies examined CHD progression in small sample sizes using Markov models, leading to imprecise estimation and limited generalizability of the findings.^48–51^ Markov models are unsuitable to describe CHD progression for two reasons. First, CHD progression is largely non-memoryless: Once a person is diagnosed with CHD-related disease, that diagnosis typically persists. Second, the Markov model is memoryless and thus does not allow for multimorbidity, a common condition that accelerates CHD progression and shortens life expectancy.^9–11^

The implication of our application study lies in two aspects. First, the estimated age-specific transition rates indicate disease mechanisms and can guide disease state-specific precision strategies targeting participants at risk of CHD. For example, compared to those without past disease history, participants with past disease history were more likely to transit from one state to another, showing the importance of disease control in the early stages of the progression. With high transition rates from heart failure to mortality observed across all ages, our study highlighted the urgency of prevention of mortality for heart failure patients starting from mid-age. Second, life expectancy in the U.S. has drastically increased over the past 50 years, primarily driven by a continuous increase in survival rate after CVD prognosis.^52, 53^ This has led to increased demand for health services and high costs for treatment and residential care. The state occupational probabilities of disease states over time can provide a more accurate estimation of disease burden and facilitate medical resources planning for real-time. Moreover, the ARIC data has unique advantages for our application, including the long-term follow-up to capture the CHD course, CHD endpoints ascertained by medical records, and access to the data through an NHLBI data repository, BioLINCC.

In summary, we developed a multi-state, non-Markov framework that allows past states to affect the transition rates of current states. The transition parameters between substates may enhance our understanding of new mechanisms of disease progression and stimulate new approaches to disease prevention. Our framework is highly suitable for estimating the progression of non-memoryless chronic diseases and has potentially broad applications in chronic disease epidemiology.

## ACKNOWLEDGMENTS

Dr. Ming Ding is supported by National Institutes of Health R21 HG012365, and Dr. Feng-Chang Lin is supported by National Center for Advancing Translational Sciences (NCATS) UM1TR004406.

## DATA AVAILABILITY STATEMENT

The R code for the non-Markov framework is available at the GitHub repository (https://github.com/mingding-hsph/A-non-Markov-regression-framework).

---

**Table S1.** Parameters used for simulation study using the ‘crisk.sim’ package in R.

| Status | Markov process with exponential distribution |  | Markov process with Weibull distribution |  | Status | Non-Markov process with exponential distribution |  | Non-Markov process with Weibull distribution |  |
| --- | --- | --- | --- | --- | --- | --- | --- | --- | --- |
|  | Ancillary parameters | beta0 | Ancillary parameters | beta0 |  | Ancillary parameters | beta0 | Ancillary parameters | beta0 |
| **Dataset 1** |  |  |  |  | **Dataset 1** |  |  |  |  |
| $S_0$ to $S_1$ | 1 | $-\log(0.2)$ | 1.5 | $-\log(0.2)$ | $S_0$ to $S_1$ | 1 | $-\log(0.2)$ | 1.5 | $-\log(0.2)$ |
| $S_0$ to $S_2$ | 1 | $-\log(0.3)$ | 1.3 | $-\log(0.3)$ | $S_0$ to $S_2$ | 1 | $-\log(0.3)$ | 1.3 | $-\log(0.3)$ |
| $S_0$ to $S_3$ | 1 | $-\log(0.3)$ | 1.5 | $-\log(0.3)$ | $S_0$ to $S_3$ | 1 | $-\log(0.3)$ | 1.5 | $-\log(0.3)$ |
| $S_0$ to $S_4$ | 1 | $-\log(0.2)$ | 1.4 | $-\log(0.2)$ | $S_0$ to $S_4$ | 1 | $-\log(0.2)$ | 1.4 | $-\log(0.2)$ |
| **Dataset 2** |  |  |  |  | **Dataset 2** |  |  |  |  |
| $S_1$ to $S_2$ | 1 | $-\log(0.4)$ | 2 | $-\log(0.4)$ | $S_1$ to $S_2$ | 1 | $-\log(0.4)$ | 2 | $-\log(0.4)$ |
| $S_1$ to $S_3$ | 1 | $-\log(0.3)$ | 1.3 | $-\log(0.3)$ | $S_1$ to $S_3$ | 1 | $-\log(0.3)$ | 1.3 | $-\log(0.3)$ |
| $S_1$ to $S_4$ | 1 | $-\log(0.3)$ | 2 | $-\log(0.3)$ | $S_1$ to $S_4$ | 1 | $-\log(0.3)$ | 2 | $-\log(0.3)$ |
| **Dataset 3** |  |  |  |  | **Dataset 3.1** |  |  |  |  |
| $S_2$ to $S_3$ | 1 | $-\log(0.5)$ | 0.9 | $-\log(0.3)$ | $S_{2\vert 0}$ to $S_3$ | 1 | $-\log(0.1)$ | 0.3 | $-\log(0.1)$ |
| $S_2$ to $S_4$ | 1 | $-\log(0.5)$ | 0.9 | $-\log(0.3)$ | $S_{2\vert 0}$ to $S_4$ | 1 | $-\log(0.2)$ | 0.5 | $-\log(0.2)$ |
|  |  |  |  |  | **Dataset 3.2** |  |  |  |  |
| | | | | | $S_{2\vert 0\_1}$ to $S_3$ | 1 | $-\log(0.5)$ | 1.2 | $-\log(0.5)$ |
| | | | | | $S_{2\vert 0\_1}$ to $S_4$ | 1 | $-\log(0.6)$ | 1.4 | $-\log(0.6)$ |
| **Dataset 4** |  |  |  |  |  |  |  |  |  |
| $S_3$ to $S_4$ | 1 | $-\log(0.5)$ | 1.8 | $-\log(0.4)$ | **Dataset 4.1.** $S_{3\vert 0}$ to $S_4$ | 1 | $-\log(0.2)$ | 0.5 | $-\log(0.2)$ |
| | | | | | **Dataset 4.2.** $S_{3\vert 0\_1}$ to $S_4$ | 1 | $-\log(0.3)$ | 1.2 | $-\log(0.3)$ |
| | | | | | **Dataset 4.3.** $S_{3\vert 0\_2}$ to $S_4$ | 1 | $-\log(0.6)$ | 2 | $-\log(0.6)$ |
| | | | | | **Dataset 4.4.** $S_{3\vert 0\_1\_2}$ to $S_4$ | 1 | $-\log(0.9)$ | 3 | $-\log(0.9)$ |

